# Formal and informal mental health support in young adults with recurrently depressed parents

**DOI:** 10.1101/2024.12.20.24319424

**Authors:** Rhys Bevan Jones, Bryony Weavers, Tessa Lomax, Emma Meilak, Olga Eyre, Victoria Powell, Becky Mars, Frances Rice

## Abstract

**Background:** A family history of mental illness, particularly parental depression, is a risk factor for mental health difficulties in young people, with this heightened risk extending into adulthood. Evidence suggests low rates of formal mental health support in children/adolescents with depressed parents, but it is unknown whether this pattern persists into adulthood and applies to informal support.

**Aims:** We examined the prevalence of formal and informal mental health support accessed by young adults with recurrently depressed parents. We identified factors associated with access to different support, reported satisfaction with support and identified potential facilitators/barriers to access.

**Methods:** A mixed-method study comprising 144 young adults (mean age=23 years, range=18-28 years) who completed psychiatric assessments and reported on their use of mental health support. Regression analyses explored predictors for support. A focus group examined facilitators and barriers.

**Results:** Young adults accessed a range of formal (29%) and informal (56%) support. Among those with psychiatric disorder, nearly half had not accessed formal support and one-fifth had not accessed any support. Predictors of support included psychiatric disorder, severity indicators (e.g. self-harm/suicidal thoughts, impairment), and demographic factors (e.g. education, gender).

Predictors varied by type of support. Most participants reported satisfaction with support. Facilitators included role models, public mental health discussions, and practitioner training. Barriers included identifying difficulties, stigma, service limitations, and family/friends’ experiences.

**Conclusions:** Young adults at high risk of mental disorders accessed various mental health support. However, many did not access/receive support when needed. Further work is required to improve access to tailored support.

## Introduction

Young people whose parents experience depression represent a recognised high-risk group for mental health difficulties^1^, most commonly depression and anxiety^2^, and the period of increased risk extends into early adult life^3^. Previous research suggests low rates of use of formal support (e.g. health services) in this population as children and adolescents^4^, but it is unknown whether this pattern continues into adult life. Indeed, young adult life is the peak period of onset for many mental health difficulties^5,6^, including in the adult children of parents with recurrent depression^3^. Moreover, there is increasing interest in young adult mental health in a clinical and research context, especially in ensuring access to support and in the development of appropriate services and resources^7^. Studies to date have also focused primarily on formal health service use in this population. Given the pressure on formal services, there is greater appreciation of the importance of informal forms of mental health support, such as self-help, online approaches, and social networks^8–10^.

It is well established that there is a significant treatment gap for mental health difficulties, including in young adults, influenced by demographic factors such as socioeconomic patterning and lower service use in young males^10–15^. Predictors of access to support include the presence of co-morbidity, self-harm/suicidal thoughts, severity of illness and impairment^13–17^. Several potential facilitators and barriers to access have been identified including individual, societal, and service/support-related factors^11,12,18,19^. A better understanding of the patterns of use of the range of services, resources, and social networks, alongside the facilitators and barriers to accessing support is important, as this could inform strategies to improve access for young adults at risk of mental health difficulties.

### Aims

This study focuses on a sample of young adults whose parents had been treated in primary care for depression. The aims were to:

- Examine the prevalence of access to support for mental health difficulties in young adults;
- Describe the types of support (e.g. services, resources, social networks) accessed and identify factors associated with use of support;
- Explore satisfaction with services and potential facilitators and barriers to accessing support.

## Methods

### Participants

The sample includes young adults from the Early Prediction of Adolescent Depression (EPAD) study, a prospective longitudinal study of the children (born between 1990 and 1998) of parents with recurrent depression^3,20–23^. The baseline sample included 337 parents (315 mothers, 22 fathers) and their biological children (aged 9-17 years (mean[SD] 12.4[2.0] years), 197 females and 140 males).

Parents and offspring were assessed separately via interview and completed questionnaires at four time points between April 2007 and September 2020. This paper focuses on the data from the fourth wave of collection which took part on average 10 years after baseline and included 197 participants, of whom 144 young adults took part in an interview and provided data on support accessed. This included 89 females and 55 males, with an age range of 18-28 (Mean=23.5 years; SD=2.30 years). Most of the sample (n=137, 95%) had two British parents, and seven had mixed (n=2) or unknown (n=5) ethnic background.

The authors assert that all procedures contributing to this work comply with the ethical standards of the relevant national and institutional committees on human experimentation and with the Helsinki Declaration of 1975, as revised in 2013. The study was approved by the Multi-Centre Research Ethics Committee for Wales (reference 06/MRE09/48) and the School of Medicine Ethics Committee, Cardiff University (reference 18/12). Written informed consent was obtained.

### Procedure

Participants were recruited primarily from general practices in south Wales. At the time of recruitment, parents were screened over the telephone to ensure they met the inclusion criteria: a history of at least two episodes of depression (DSM-IV Major Depressive Disorder, MDD^24^) later confirmed at baseline using diagnostic interview and had a biologically related child living at home aged 9-17 years. Families were excluded if the parent had a diagnosis of bipolar or psychotic disorder at baseline or if the child had a moderate to severe learning disability (IQ<50). If there was more than one eligible child in the household, the youngest child was selected for participation.

Most assessments took place in the participant’s home with young adults and parents interviewed separately. A small number of assessments were undertaken over the telephone/video call as required.

### Measures

#### Mental health support

Participants were given a list of support sources and asked to indicate (yes/no) whether they were ‘currently seeing or using’ any of these for help with mental health issues. They could also provide a free-text response under ‘someone else’ if not listed. Data were categorised into the binary variables ‘formal support’ and ‘informal support’. Formal support included primary care (general practitioner), secondary care (mental health specialist: psychiatrist, clinical psychologist, mental health nurse), or other formal support (e.g. counsellor, social services, student support). Informal support included self-help (e.g. internet-based therapy/apps, self-help group), internet use (for information or advice), or family member or close friend.

Participants were asked whether they were satisfied with the help received if they had ‘ever used services for help with mental health’ (options: yes, no, N/A) and why (free-text response). This was asked about help from services in general, and not for each type of support individually.

#### Predictors of support

Predictors were selected based on prior literature^10–17^.

##### Current psychiatric diagnoses

These were assessed using a semi-structured diagnostic interview, the Young Adult Psychiatric Assessment (YAPA)^25^. The YAPA was used in separate interviews with parents and young adults to assess offspring DSM-IV^24^ psychopathology in the preceding 3 months. The parent interviews asked about symptoms of depression and attention deficit hyperactivity disorder (ADHD) in their offspring, whereas the young adult interviews included assessment of a wide range of psychiatric disorders. Cases where the young adults met criteria for a psychiatric disorder or had subthreshold symptoms were reviewed by two psychiatrists and diagnoses were agreed by clinical consensus.

Three variables were considered as predictors of service use: a diagnosis of any psychiatric disorder, a depressive disorder and an anxiety disorder, as these are the most common disorders in this population^3^.

A diagnosis of ‘any psychiatric disorder’ included depressive disorders (MDD, dysthymia, cyclothymia, and adjustment disorder), anxiety disorder (generalised anxiety disorder (GAD), social anxiety, separation anxiety, agoraphobia, obsessive-compulsive disorder, and panic disorder), ADHD, conduct disorder, and personality disorders (schizotypal and borderline). Although personality disorders were not explicitly assessed by the standardized interview used, in a small number of cases, a personality disorder diagnosis was judged by clinical consensus to be appropriate for the symptoms exhibited.

##### Comorbidity

This was defined as those currently meeting diagnostic criteria for two or more DSM-IV disorders and categorised as a binary variable (yes/no).

##### Self-harm or suicidal thoughts

The presence of self-harm/suicidal thoughts was assessed using the YAPA over the last 3 months. Responses to these questions were combined and categorised as a binary variable (yes/no).

##### Total difficulties and impairment

Measures of total difficulties (total score, continuous) and impairment (impairment score, continuous), associated with emotional or behavioural problems, were indicated by the self-report Strengths and Difficulties Questionnaire (SDQ) impact supplement^26^.

##### Demographic factors

These included: i) gender (female/male/other), ii) age in years, iii) poor social support (i.e. only one person or no-one to rely on), iv) living alone, v) NEET (not in education, employment or training), vi) education status (not completed degree and not currently in university), and vii) low personal income (categorised as below £18,000/annum^27^).

For further details on the measures, see Supplement 1.

### Focus group

A focus group session was held with young adults from the EPAD study and a member of the National Centre for Mental Health youth advisory group via videoconferencing to explore access to various types of support, and to enrich the quantitative findings. The group was facilitated by RBJ, EM and a research assistant, following a pre-prepared topic guide (Supplement 2). The session was held in 2021 and lasted approximately 90 minutes. Mentimeter was used to gather answers to specific questions and to encourage discussion. We aimed for a balance in terms of the gender and age of participants. The focus group was digitally audio-recorded and transcribed; participants could also contribute through the videoconferencing platform’s ‘chat function’ or by emailing researchers separately.

### Analysis

#### Statistical analysis

We first describe the sample characteristics (proportions or means as appropriate) in the whole sample. Next, we describe the proportions of participants using different types of support in the whole sample and separately by psychiatric disorder status, because those meeting criteria for a disorder are more likely to require support.

A series of univariable logistic regression analyses were then conducted to investigate predictors of the three support outcome variables (any formal support, any informal support, any support (formal and informal combined)), firstly in the whole sample, and then in the subsample with psychiatric disorder. Analyses reported in main text use inverse probability weights (IPW)^28^ to account for attrition between study baseline and the fourth follow-up phase, the focus of this analysis. IPW were calculated by examining variables at the baseline assessment that predicted missingness from the analysis sample consistent with previous publications^3^ (Supplement 3). Tables report results using IPW. Results were broadly similar when analysing complete cases and IPW (Supplement 4). A sensitivity analysis was conducted excluding family/friend support from the informal support category, to examine sources developed to provide self-guided support and because social support was included as a predictor. Data on satisfaction with services was presented descriptively (percentages). Statistical analysis was conducted using SPSS (version 26, IBM).

#### Qualitative analysis

The focus group transcript was analysed using a thematic analysis approach. This is a process of identifying, analysing, reporting, and interpreting patterns or themes^29^. To ensure the reliability of coding, the transcript was coded by RBJ and double-coded independently by EM. Initial ideas on the coding framework were discussed among the team; the draft framework was applied to some of the data and refined as coding proceeded. Codes were applied to broad themes, which were then broken down further into subcodes. Transcripts were examined to identify the key themes and associated subthemes. A similar analysis approach was taken with the free-text responses regarding satisfaction with services.

## Results

### Prevalence of mental health difficulties and demographic factors

Table 1 shows the psychiatric and demographic characteristics of the sample. Over a third (38.7%) of individuals in the sample met criteria for a current psychiatric disorder, with 24.7% having a depressive disorder and 25.2% an anxiety disorder. Comorbidity was identified in 17.2% of individuals and 12.4% had recent self-harm/suicidal thoughts. A quarter of the sample (24.7%) reported poor social support, and 13.1% were living alone. Sixteen percent (16.1%) were NEET and 43.1% had not completed a degree and were not currently in university. Over two-thirds (68.5%) had a personal annual income under £18,000.

**Table 1.** Prevalence of mental health difficulties and demographic factors (IPW applied)

| Mental health difficulties | Full sample,<br>% or mean (SD)<br>N=144 |
| --- | --- |
| Any current psychiatric disorder | 38.7 |
| Any current depressive disorder | 24.7 |
| Any current anxiety disorder | 25.2 |
| Current comorbidity | 17.2 |
| Current self-harm or suicidal thoughts | 12.4 |
| SDQ total difficulties score | 11.7 (5.99) |
| SDQ impairment score | 1.5 (2.25) |
| Social, educational, and occupational factors |  |
| Gender (female) | 61.8 |
| Age (years) | 23.5 (2.30) |
| Poor social support | 24.7 |
| Living alone | 13.1 |
| Not in education, employment, or training (NEET) | 16.1 |
| Not completed degree and not currently in university | 43.1 |
| Personal income <£18,000 per annum | 68.5 |

### Prevalence of access to support

Table 2 provides information about the use of different types of support. Among the whole sample, 60.2% of individuals reported currently receiving some form of mental health support. Informal support was used by a greater proportion than formal support (55.9% vs 29.3%). With regards to formal support, primary care was most frequently used (23.0%), followed by secondary care (10.3%) and other formal support (7.2%). For informal support, family/friends were the most reported (55.9%), followed by the internet (19.1%) and self-help (4.4%). When excluding family/friend support, the proportion accessing informal support reduced to 22.3%.

**Table 2:** Support accessed for mental health difficulties in the whole sample, and in those with and without a current psychiatric disorder (IPW applied)

| Type of support accessed | Whole sample<br>%<br>N=144 | Any psychiatric disorder<br>%<br>N=53 | No psychiatric disorder<br>%<br>N=91 |
| --- | --- | --- | --- |
| **Formal support** |  |  |  |
| **Medical support** |  |  |  |
| Primary Care | 23.0 | 44.8 | 9.1 |
| Secondary Care | 10.3 | 21.4 | 3.1 |
| Medical support total | 27.3 | 52.9 | 11.2 |
| **Other formal support** | 7.2 | 13.7 | 3.0 |
| **Any formal support (total)** | 29.3 | 56.5 | 12.1 |
| **Informal support** |  |  |  |
| **Self-guided support** |  |  |  |
| Self-help | 4.4 | 9.7 | 1.1 |
| Internet | 19.1 | 31.4 | 11.4 |
| Self-guided support total | 22.3 | 37.9 | 12.5 |
| **Family member or close friend** | 55.9 | 72.0 | 43.4 |
| **Any informal support (total)** | 55.9 | 74.3 | 44.3 |
| **Any support total**<br>**(formal or informal)** | 60.2 | 82.0 | 46.4 |
| **Any support (excluding family**<br>**member/friend)** | 39.3 | 68.7 | 20.7 |

**Footnote for Table 2:** Any formal support (total): Primary care, secondary care, or other formal support. Any informal support (total): Self-help, Internet or family member/close friend. Any support total: Any formal or informal support.

Access to support was higher among those with disorder compared to those without (formal 56.5% vs 12.1%; informal 74.3% vs 44.3%; any support 82.0% vs 46.4%). However, 43.5% of those with disorder were not in contact with formal services and 18.0% did not receive any support, or 31.3% when excluding family/friend support. Among those with disorder, family/friends were the most reported type of support (72.0%), followed by primary care (44.8%) and the internet (31.4%). Again, the proportion of those with disorder accessing informal support was lower (37.9%) when discounting family/friend support.

### Predictors of support in the whole sample

Findings from the regression analyses in the whole sample are shown in Table 3. Several variables were consistently associated with both formal and informal support. These included the presence of any psychiatric disorder, depressive disorder, anxiety disorder, self-harm/suicidal thoughts, and higher SDQ difficulties and impairment scores. For education status, those without a degree and not in university were less likely to access formal support, and there was weak evidence for a negative association with informal support. There was weak evidence for an association with gender, with females slightly more likely to access both types of support than males. The odds ratios (ORs) were often higher for formal than informal support, particularly for the disorder categories, although there was some overlap in the confidence intervals.

**Table 3:** Regression analysis on current support accessed by young adults in the whole sample (N=144) (IPW applied)

|  | Any formal support |  | Any informal support |  | Any support |  |
| --- | --- | --- | --- | --- | --- | --- |
|  | OR<br>(95% CI) | P-value | OR<br>(95% CI) | P-value | OR<br>(95% CI) | P-value |
| **Any current psychiatric disorder** | 9.4 (5.46 – 16.12) | <.001 | 3.6 (2.25 – 5.85) | <.001 | 5.3 (3.13 – 8.91) | <.001 |
| **Any current depressive disorder** | 5.9 (3.46 – 10.09) | <.001 | 2.2 (1.30 – 3.74) | .003 | 4.1 (2.20 – 7.47) | <.001 |
| **Any current anxiety disorder** | 6.4 (3.77 – 10.99) | <.001 | 4.8 (2.65 – 8.74) | <.001 | 5.5 (2.86 – 10.47) | <.001 |
| **Current comorbidity** | 7.0 (3.77 – 12.85) | <.001 | 1.5 (0.85 – 2.76) | .15 | 3.6 (1.77 – 7.32) | <.001 |
| **Current self-harm/suicidal thoughts** | 3.5 (1.80 - 6.78) | <.001 | 3.19 (1.48 - 6.88) | .003 | 2.6 (1.22 – 5.65) | .014 |
| **SDQ total difficulties score** | 1.2 (1.12 – 1.23) | <.001 | 1.1 (1.06 – 1.15) | <.001 | 1.1 (1.09 – 1.20) | <.001 |
| **SDQ impairment score** | 1.6 (1.39 – 1.84) | <.001 | 1.4 (1.18 – 1.55) | <.001 | 1.4 (1.21 – 1.64) | <.001 |
| **Gender (female)** | 1.5 (0.94 – 2.51) | .09 | 1.5 (0.96 – 2.31) | .08 | 1.5 (0.93 – 2.25) | .10 |
| **Age (years)** | 0.95 (0.85-1.05) | .29 | 0.98 (0.89-1.07) | 0.61 | 0.99 (0.90-1.09) | .87 |
| **Poor social support** | 1.2 (0.69 – 2.11) | .51 | 2.2 (1.33 – 3.66) | .002 | 1.6 (0.95 – 2.59) | .08 |
| **Living alone** | 2.7 (1.36 – 5.22) | .004 | 0.8 (0.44 – 1.63) | .61 | 2.1 (0.98 – 4.34) | .06 |
| **Not in education, employment, or training (NEET)** | 3.6 (1.96 – 6.79) | <.001 | 1.2 (0.65 – 2.23) | .56 | 2.1 (1.05 – 4.05) | .04 |
| **Not completed degree and not currently in university** | 0.4 (0.23 – 0.61) | <.001 | 0.7 (0.43 – 1.03) | .07 | 0.5 (0.31 – 0.76) | .002 |
| **Personal income <£18,000 per annum** | 1.3 (0.66 – 2.41) | .49 | 1.7 (0.97 – 2.90) | .06 | 1.9 (1.08 – 3.24) | .03 |

Comorbidity, living alone, and NEET were associated only with formal support whereas poor social support was only associated with informal support. There was weak evidence for an association between low personal income and informal support. Age was the only variable that was not associated with either type of support. In sensitivity analysis (Supplement 5) excluding family/friend support from the informal support outcome, there were associations with comorbidity and NEET in addition to those noted above. However, associations were no longer found with self-harm/suicidal thoughts, gender and education status (previously weak evidence).

### Predictors of support in young adults with psychiatric disorder

Findings from the regression analyses among those with disorder are shown in Table 4. Those with comorbidity and greater SDQ total difficulties and impairment scores were more likely to access formal support. Those without a degree and not in university and those with a low personal income were less likely to access formal support.

**Table 4:** Regression analysis on current support accessed by young adults with a psychiatric disorder (N=53) (IPW applied)

|  | Any formal support |  | Any informal support |  | Any support |  |
| --- | --- | --- | --- | --- | --- | --- |
|  | OR<br>(95% CI) | P-value | OR<br>(95% CI) | P-value | OR<br>(95% CI) | P-value |
| **Current comorbidity** | 2.02 (0.99-4.12) | .05 | 0.39 (0.18-0.88) | .02 | 0.99 (0.40-2.42) | .97 |
| **Current self-harm/suicidal thoughts** | 0.69 (0.39-1.86) | .69 | 0.93 (0.40-2.32) | .16 | 0.49 (0.19-1.26) | 0.14 |
| **SDQ total difficulties score** | 1.15 (1.07-1.25) | <.001 | 1.02 (0.95-1.10) | .64 | 1.09 (1.00-1.19) | .05 |
| **SDQ impairment score** | 1.41 (1.18-1.69) | <.001 | 1.18 (0.99-1.41) | .06 | 1.16 (0.96-1.40) | .13 |
| **Gender (female)** | 0.89 (0.44-1.79) | .73 | 1.80 (0.82-3.97) | .15 | 1.26 (0.51-3.09) | .62 |
| **Age (years)** | 0.94 (0.81-1.09) | .40 | 0.87 (0.73-1.02) | .09 | 0.85 (0.70-1.03) | .10 |
| **Poor social support** | 1.88 (0.85-4.16) | .12 | 4.24 (1.81-9.94) | <.001 | 2.39 (0.93-6.11) | .07 |
| **Living alone** | 2.42 (0.88-6.65) | .09 | 0.44 (0.16-1.17) | .10 | 2.89 (0.61-13.63) | .18 |
| **Not in education, employment, or training (NEET)** | 1.48 (0.68-3.20) | .33 | 0.20 (0.08-0.48) | <.001 | 0.43 (0.17-1.07) | .07 |
| **Not completed degree and not currently in university** | 0.34 (0.17-0.71) | .004 | 0.62 (0.28-1.36) | .23 | 0.16 (0.06-0.48) | .001 |
| **Personal income <£18,000 per annum** | 0.32 (0.11-0.88) | .03 | 2.61 (0.83-8.21) | .10 | 2.61 (0.83-8.21) | .10 |

Those with comorbidity and who were NEET were less likely to access informal support. There was weak evidence for an association with SDQ impairment and a negative association with age. Those with poor social support were more likely to access informal support, and this association remained (although weaker) after sensitivity analysis when excluding family/friends from informal support (Supplement 5). A similar pattern of sensitivity analysis results was found for SDQ impairment and age, although associations were no longer found for comorbidity or NEET. Additional associations were found – those with self-harm/suicidal thoughts and living alone were less likely to access support and those with higher SDQ difficulties scores were more likely.

### Satisfaction with services

Ninety-three young adults (64.6% of the whole sample) reported having ever used services for help with mental health and provided information on satisfaction with services. Of these, over two-thirds (69.6%) were satisfied with the help received and 21.5% were not satisfied. The remainder answered both ‘Yes’ and ‘No’ (6.5%) or ‘don’t know’ (2.2%).

The most common reasons given for satisfaction with services (Supplement 6) included being taken seriously, feeling listened to and understood, being helped to rationalise, talking to someone impartial, and speed of appointment. Reasons for dissatisfaction included long waiting times, disjointed services, feeling dismissed/unsupported, being offered medication too quickly, and poor relationships with professionals.

### Qualitative (focus group) results

Six people agreed to participate in the focus group, however, two did not attend. Of the four participants, two identified as female, one as male, and one as non-binary. One was aged 21-23 and three were aged 24-27. Three were working full-time and one was in full-time education. Three had experienced mental health difficulties, and all had sought support/advice (e.g. health services, helplines, websites) for such difficulties.

The themes, subthemes and verbatim examples are presented in Supplement 7. The key themes were:

1) young adults access a range of online and informal support (e.g. internet resources, virtual sessions, apps, charities, trusted people);
2) facilitators for help-seeking (e.g. role models, ‘talking’ about mental health, practitioner education, emphasis on psychological approaches);
3) difficulties with identifying one’s mental health problems as a barrier to accessing help, with subthemes including conveying feelings, acknowledging and minimising difficulties, and changes as people get older;
4) societal and service/support-related factors as barriers, with subthemes concerning lack of awareness of help, stigma associated with health services, service pressures, and concerns about biases;
5) effects of seeing how someone close to you experienced and managed their problems and experienced health services and other help.

To address barriers to accessing support and help-seeking, participants suggested increasing the number of role models for young adults, providing clear guidance on support pathways, promoting a ‘positive narrative’ around mental health, and developing educational and self-help resources (e.g. through charities and health services).

## Discussion

This paper examined access to formal and informal types of mental health support in a sample of young adults at high risk of mental health difficulties due to a family history of recurrent depression. Sixty percent across the whole sample reported access to some form of support and 29% used formal support. Access to formal and informal support was higher amongst those with a psychiatric disorder, however 44% of this group were not in contact with any formal services, and just under 1 in 5 received no support at all. This figure rises to about 1 in 3 when excluding social networks.

A wide range of formal and informal support were accessed. Access to support among the whole sample was predicted by diagnosis as well as other indicators of severity of difficulties (e.g. self-harm/suicidal thoughts, SDQ scores). Associations were consistently found for demographic predictors such as education status and gender. There were however some differences found for the remaining factors according to the type of support (formal/informal) and in the subsample with disorder. Over two-thirds were satisfied with the help received from services. These findings, together with the results from the focus group identified several barriers and facilitators to accessing support. Of relevance to this group of young adults with depressed parents, was that their help-seeking behaviour could be influenced by how those close to them (e.g. parents/carers) managed their health difficulties and their experiences of accessing support.

### Comparison with existing literature

Service use has been previously examined in this cohort when the participants were aged 9-17 years^4^. At that time, only a third of those with psychiatric disorder were in contact with services. However, only formal service use was examined, including educational, social, youth justice and health services. The current study builds on this work by including informal services, and with the focus on early adult life, a developmental transition to independence associated with the emergence of mental health difficulties^5,6^, changes in support services, and personal and social changes and challenges (e.g. education, employment, relationships)^30^. The current work also helps to address the lack of long-term studies in this population and suggests that access to formal support among those with disorder increases from childhood/adolescence to young adulthood (from one-third to just over half the sample).

The levels of support accessed in this sample are higher than reported in some earlier studies involving young adults with mental health difficulties in the UK. Salaheddin et al showed that 65% of 16-25-year-olds with mental health difficulties accessed formal or informal help (including peer support)^11^. This is compared to 82% of those with disorder who accessed any support in our study. This might suggest that individuals with a parent (known to services) with mental health difficulties may be more likely to seek support, or this might be explained by differences in methodology (e.g. participant characteristics, definition of difficulties and support).

Factors associated with access to support in the current study are consistent with those found in previous studies of young adults in the UK such as mood disorders, severity of difficulties, comorbidity, suicide risk and female gender^12,14,16^. The current study extends this work by looking at a wide range of sociodemographic factors (including age, social support, living alone, NEET, education status, and personal income) and suggests that for some of these factors, associations may be different for formal and informal support. The barriers to access identified in our focus group are also consistent with those arising from the literature, including those related to stigma^11,13,18,19^, difficulty in identifying or expressing concerns^11,17,19^ and being unsure where to go^13^. Particularly relevant to this work, an earlier review concluded that stigma related to families with parental mental illness can prevent family members from seeking support^31^.

### Strengths and limitations

Data were drawn from a large study of young adults at elevated risk for psychopathology, recruited mainly from general practice, and followed prospectively over 13 years (from childhood/adolescence) and across key developmental phases. Assessments were rigorous, involving multiple informants and diagnostic interviews, and access to a broad range of support was considered.

The findings must however be interpreted considering the following limitations. The interview used to capture mental health support relied on the individual’s recall and interpretation of the question on whether they were currently receiving/using support. Therefore, some forms of support may have gone unreported. Broad definitions for support were used, although sensitivity analyses were conducted excluding social networks from informal support. Data on satisfaction with help received were based on lifetime reporting and was not specific to the type of services accessed. As with all longitudinal studies, there was some attrition. Of the original 337 families in the sample, 197 (58.5%) took part in wave 4, with 144 participants having data on both disorder and support, leading to small subsamples for certain analyses (e.g. those with disorder). However, IPW was used to account for attrition.

### Implications for practice

Young adults in this study accessed a range of support, and distinct levels and types of support are likely to be required based on individual needs and severity of difficulties/impairment. UK guidelines for depression recommend tailored approaches including guided self-help, psychotherapy, and medication, depending on the presentation from subthreshold/mild to severe depression^32^. A UK study of older adolescents^33^ found beneficial treatment effects on depressive symptoms only in those who met criteria for psychiatric disorder or had high subthreshold symptoms and impairment, suggesting that this is a suitable threshold for formal services.

In this sample, 44% of young adults with disorder were not accessing any formal support suggesting that many in need of support are not receiving it. Access to support among this subgroup was predicted by indicators of severity and impairment alongside demographic factors such as living alone. Specifically, those with lower education status and personal income were less likely to seek formal support, suggesting they represent a hard-to-reach group where targeted interventions could improve access. Education status was also negatively associated with formal support in the whole sample, while those who were NEET were more likely to access support. These findings may appear contradictory but could be explained by higher levels of difficulties and impairment among those who are unemployed.

The proportion of participants with disorder receiving support increased from 56.5% (formal support) to 82% when including informal support, highlighting the reliance on less formal sources. This can be explained in part by the lack of formal services, the growth in self-help, online resources, and digital devices, as well as young adults’ comfort with informal support, attributable to factors like accessibility, convenience, trust, confidentiality, and stigma^10,34^. This also reflects the importance of social networks, and the proportion accessing any support reduced to 68.7% after excluding family/friends.

Whilst focus group participants suggested that young adults are increasingly open about mental health and seeking help, they also highlighted barriers to access including difficulties identifying symptoms, awareness of support, and prejudices related to services. Their recommendations for educational and anti-stigma programmes align with the call for help-seeking interventions to improve mental health knowledge and stigma^34^. Participants asked for better coordination among health and student services and charities. Acceptable and effective resources/services are needed, that are co-developed with users^35^.

### Conclusions

All participants in this study were young adults with parents with recurrent depression and were therefore at elevated risk of mental health difficulties. Participants accessed a variety of sources of mental health support, with just over half of those with a psychiatric disorder accessing formal help, and one in five were not receiving any support. Further work is needed to ensure early identification of difficulties and access to support, and a better understanding of the types of support that meet the needs and preferences of young adults, including those at risk.

**Rhys Bevan Jones** Wolfson Centre for Young People’s Mental Health/Division of Psychological Medicine and Clinical Neurosciences, Cardiff University & Cwm Taf Morgannwg University Health Board, Wales; **Bryony Weavers** Wolfson Centre for Young People’s Mental Health/Division of Psychological Medicine and Clinical Neurosciences, Cardiff University, Wales; **Tessa Lomax** Oxford Health NHS Foundation Trust and Department of psychiatry, University of Oxford, Warneford Hospital, Oxford, England; Wolfson Centre for Young People’s Mental Health/Division of Psychological Medicine and Clinical Neurosciences, Cardiff University & Cwm Taf Morgannwg University Health Board, Wales; **Emma Meilak** Wolfson Centre for Young People’s Mental Health/Division of Psychological Medicine and Clinical Neurosciences, Cardiff University, Wales; **Olga Eyre** Wolfson Centre for Young People’s Mental Health/Division of Psychological Medicine and Clinical Neurosciences, Cardiff University, Wales; **Victoria Powell** Wolfson Centre for Young People’s Mental Health/Division of Psychological Medicine and Clinical Neurosciences, Cardiff University, Wales; **Becky Mars** Centre of Academic Mental Health, Population Health Sciences, University of Bristol & NIHR Bristol Biomedical Research Centre, University Hospitals Bristol and Weston NHS Foundation Trust and University of Bristol, England; **Frances Rice** Wolfson Centre for Young People’s Mental Health/Division of Psychological Medicine and Clinical Neurosciences, Cardiff University, Wales.

### Supplementary material

1) Measures for support, functioning and impairment,
2) Topic guide for focus group,
3) Missing data and IPW,
4) Results from regression analysis on support accessed, without IPW,
5) Results from sensitivity analysis without family/friends support, with IPW,
6) Qualitative responses on satisfaction with services,
7) Themes, subthemes, and quotes from focus group.

## Acknowledgements

The authors are grateful to all the participating families in the EPAD study. The authors thank the GPs and psychiatrists who helped with this study, including Dr Robert Potter. The authors thank all the assistant psychologists and research assistants who carried out data collection and helped with the focus group, including Jessica Lennon and Alice Stephens.

## Author contributions

Conceptualization RBJ, FR. Preparation of manuscript RBJ. Statistical analysis BW. Literature review TL, RBJ. Qualitative analysis RBJ, EM. Funding acquisition and supervision FR. Writing-review & editing – all authors. All authors approved the final draft.

## Data availability

Due to ethical restrictions, data collected at assessment waves 1 to 3 cannot be made openly available. Supporting data collected at assessment wave 4 is openly available from the Cardiff University data repository at http://doi.org/10.17035/d.2023.0263728184.

## Tables – Formal and informal mental health support in young adults with recurrently depressed parents

## Supplementary Materials

### Supplement 1: Measures for support, difficulties, functioning and impairment outcomes in early adult life

#### Mental health support

Formal support included primary care (general practitioner), secondary care (mental health specialist: psychiatrist, clinical psychologist, mental health nurse), or other formal support (counsellor, social services, student support services, advocate, call line, support worker, wellbeing team, private sleep therapy, hypnotherapist, other therapy). Informal support included self-help (internet-based therapy, self-help group, italk, online meditation, Headspace app, NHS self-help, MoodGYM), internet use (for information or advice), or family member or close friend.

#### Psychiatric diagnoses

For ADHD and MDD, a diagnosis was present if reported by either the parent or the young adult, as had been done in previous waves with this cohort. Parent and child reports were highly correlated.

#### Self-harm/suicidal thoughts

As part of the YAPA during wave 4 interviews, the young adults reported whether they wanted to die, tried to hurt, or kill themselves, thought that life was not worth living, wished they were dead or done anything that made people think that they wanted to die.

#### Distress and impairment

Young adult and parent-reports on the impact supplement of the Strengths and Difficulties Questionnaire (SDQ) were used to assess distress and impairment (at home, school, in friendships or in leisure activities) associated with mental health problems. Five items with responses of “Not at all” (0), “Only a little” (0), “A medium amount” (1) or “A great deal” (2) were summed to give a maximum total score of 10. Those scoring 1 were classed “borderline” and those scoring 2 or more were classed as “abnormal” as recommended previously. Child and parent-reported “borderline” or “abnormal” scores were combined using an either/or approach. Parent and child reports were highly correlated.

#### SDQ impairment score

Responses to the questions on chronicity and burden to others are not included in the impact score. When respondents have answered ‘no’ to the first question on the impact supplement (i.e. when they do not perceive themselves as having any emotional or behavioural difficulties), they are not asked to complete the questions on resultant distress or impairment; the impact score is automatically scored zero in these circumstances.

#### Social support

As part of the interview at wave 4, the young adults were asked to list the people they could most rely on for social support. From this, a binary variable was derived for those with only one or no people to rely on, versus those with two or more people they could rely on.

#### Education and employment

Young adults reported on their education and employment via questionnaire. A binary variable (0=no, 1=yes) capturing whether the young person was not currently in education, employment, or training (NEET status) was derived (NEET=Not currently in full time, part time or occasional work, doing an apprenticeship, in full-time education or self-employed. Includes those who are unemployed, unable to work due to sickness/disability or full/part-time carers). An additional binary variable for whether the young person had not completed a degree and was not currently in university was derived.

### Supplement 2: Topic guide for focus group

#### Access to mental health support in young adults

##### Introduction

- Thank them for taking part in this project.
- To explain want to record this session. The recording is only to transcribe the audio of the meeting.
- Summarise themes to discuss.
- Discuss processes & ‘ground rules’ with the participants e.g., to be respectful of each other’s comments, and to maintain confidentiality.
- Initial ‘ice breakers’ to ensure the group feel comfortable.

##### Access to mental health support & help-seeking

- Opening discussion point – any general comments about young adult’s access to support or help-seeking for mental health (MH) difficulties?
- Do you think young adults of your age go for help if they have MH difficulties?
- Where would you/they go for help? (e.g. family/peers, GP, charities, internet…)
- A common finding is that lots of young adults who are experiencing MH difficulties, do not get or seek help for their MH.
- Why do you think that might be? During pandemic and pre-pandemic?
- What barriers are there to getting/seeking help for MH?
- How could we overcome these?
- What facilitators to seeking help for MH are there?
- How do you feel about getting help from outside MH services, for example through charities? Compared to MH services? Advantages? Disadvantages?
- How do you feel about using internet or mobile phone resources for MH difficulties? Compared to face-to-face? Advantages? Disadvantages?
- Do you think that having someone in the family (or someone close to you) with MH difficulties affects whether someone might go for help for their own MH? (links with EPAD sample)
- Do you think that where/how people go for help change as they get older e.g. from teenage years to young adulthood? (links with progression from waves in EPAD)

##### Final comments

Any final comments on the programme or study?

Finish on something relaxing or upbeat to ensure participants not ruminating over what we’ve talked about. Before we finish, could everyone say one thing they are going to do to practice self-care or make them feel good after this meeting or over the next day or so?

Thank them for participating. At the end of the meeting, we will also remind them to contact us if required and signpost them to relevant MH resources.

### Supplement 3: Missing data and inverse probability weighting (IPW)

To account for the impact of attrition across the waves (baseline to fourth follow-up), inverse probability weighting (IPW)^28^ was used. This involved weighting the analysis sample by the inverse probability of being missing. Variables measured at baseline (wave 1) were examined as predictors of missingness at wave 4, consistent with previous publications^6^. Variables that predicted missingness at wave 4 were if the family was from a single parent household (B=1.67, p=.041), parent low educational attainment, defined as not achieving GCSE level or equivalent (B=2.39, p=.011), parental low income, defined as a household income of £20,000 or less per annum (B=1.82, p=.002). The presence of psychiatric disorder in the child at baseline (N=1.58, p=.099) was also included in the missingness model as it related to the study outcomes. Minimal missing data on indicators used to derive weights were singly imputed as the modal value (all indicators had <13% missing data). The Hosmer and Lemeshow test indicated that the model was an acceptable fit (X^2^ =2.72 (df=4), p=.607). Weights ranged from 1.94 to 6.59.

### Supplement 4: Results from Tables 1-4 in the main text, without IPW

**Table 1:**
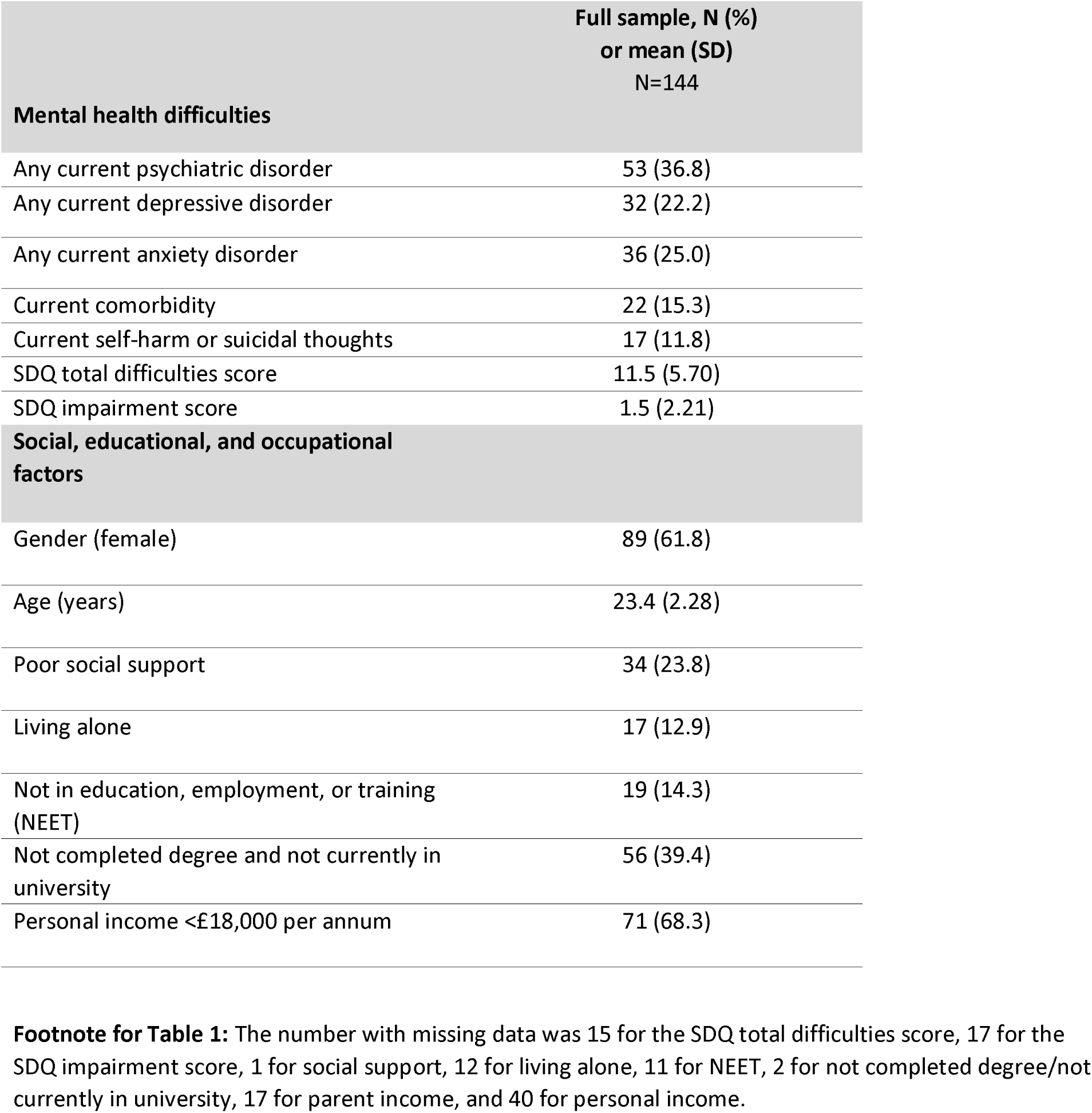
Prevalence of mental health difficulties and demographic factors.

**Table 2:**
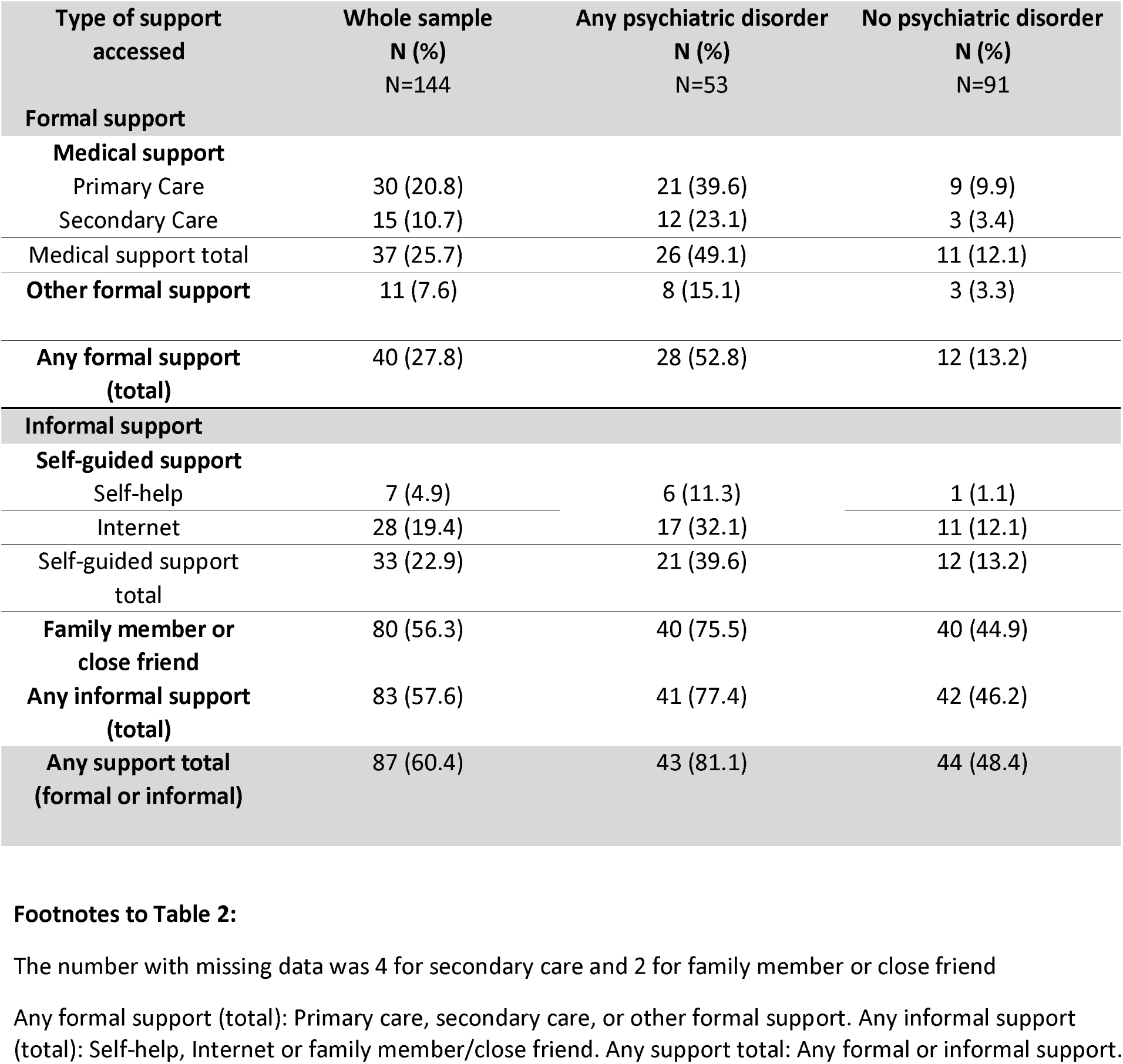
Support accessed for mental health difficulties in the whole sample, and in those with and without a current psychiatric disorder.

**Table 3:** Regression analysis on current support accessed by young adults in the whole sample (N=144)

|  | Any formal support |  | Any informal support |  | Any support |  |
| --- | --- | --- | --- | --- | --- | --- |
|  | OR<br>(95% CI) | P-value | OR<br>(95% CI) | P-value | OR<br>(95% CI) | P-value |
| **Any current psychiatric disorder** | 7.4 (3.27 – 16.61) | <.001 | 4.0 (1.86 – 8.56) | <.001 | 4.6 (2.06 – 10.24) | <.001 |
| **Any current depressive disorder** | 5.3 (2.30 – 12.31) | <.001 | 2.7 (1.10 – 6.40) | .03 | 3.6 (1.36 – 9.33) | .01 |
| **Any current anxiety disorder** | 5.5 (2.43 – 12.45) | <.001 | 4.1 (1.67 – 10.27) | <.002 | 4.5 (1.72 – 11.62) | .002 |
| **Current comorbidity** | 5.1 (1.96 – 13.16) | <.001 | 1.7 (.65 – 4.47) | .28 | 2.5 (.88 – 7.29) | .09 |
| **Current self-harm/suicidal thoughts** | 2.5 (1.15 – 5.24) | .02 | 2.2 (1.06 – 4.65) | .04 | 2.2 (1.02 – 4.61) | .04 |
| **SDQ total difficulties score** | 1.2 (1.07 – 1.25) | <.001 | 1.1 (1.03 – 1.18) | .01 | 1.1 (1.03 – 1.19) | .01 |
| **SDQ impairment score** | 1.6 (1.28 – 1.98) | <.001 | 1.4 (1.10 – 1.72) | .01 | 1.4<sup>+</sup> (1.11 – 1.80) | .01 |
| **Gender (female)** | 2.0 (.92 – 4.48) | .08 | 1.6 (.81 – 3.06) | .18 | 1.7 (.86 – 3.31) | .13 |
| **Age (years)** | 1.0 (0.85 – 1.18) | .96 | 1.0 (0.86 – 1.15) | .94 | 1.0 (0.88 – 1.19) | .74 |
| **Poor social support** | 1.7 (.66 – 4.23) | .28 | 2.1 (.96 – 4.58) | .06 | 1.8 (.82 – 3.91) | .14 |
| **Living alone** | 2.0 (.69 – 5.69) | .20 | .99 (.35 – 2.78) | .98 | 1.6 (.53 – 4.85) | .41 |
| **Not in education, employment, or training (NEET)** | 2.8 (1.02 – 7.49) | .05 | 1.6 (.56 – 4.44) | .39 | 1.9 (.64 – 5.62) | .25 |
| **Not completed degree and not currently in university** | 0.4 (.21 – .93) | .03 | 0.7 (.35 – 1.37) | .29 | 0.6 (.28 – 1.15) | .12 |
| **Personal income <£18,000 per annum** | 1.4 (.51 – 3.64) | .54 | 1.5 (.63 – 3.32) | .39 | 1.6 (.71 – 3.76) | .25 |

**Table 4:** Regression analysis on current support accessed by young adults with a psychiatric disorder (N=53)

|  | N (%) or mean (SD) | Any formal support |  | Any informal support |  | Any support |  |
| --- | --- | --- | --- | --- | --- | --- | --- |
|  |  | OR (95% CI) | P-value | OR (95% CI) | P-value | OR (95% CI) | P-value |
| **Current comorbidity** | 22 (41.5) | 1.5 (.51 – 4.5) | .44 | 0.4 (.11 – 1.53) | .19 | 0.7 (.16 – 2.61) | .55 |
| **Current self-harm/suicidal thoughts** | 30 (56.6) | 0.8 (.29 – 2.29) | .64 | 1.4 (.39 – 5.13) | .60 | 0.8 (.21 – 3.32) | .81 |
| **SDQ total difficulties score** | 15.1 (5.53) | 1.1 (1.00 – 1.28) | .05 | 1.0 (.90 – 1.15) | .82 | 1.0 (.91 – 1.19) | .53 |
| **SDQ impairment score** | 2.9 (2.80) | 1.5 (1.10 – 1.96) | .01 | 1.2 (.90 – 1.64) | .20 | 1.2 (.89 – 1.66) | .22 |
| **Gender (female)** | 33 (62.3) | 1.2 (.39-3.65) | .75 | 1.9 (.52 – 7.10) | .32 | 1.9 (.47 – 7.49) | .38 |
| **Age (years)** | 23.1 (2.26) | 1.0 (0.80 – 1.30) | .86 | 0.9 (0.69 – 1.23) | .57 | 1.0 (0.70 – 1.29) | .95 |
| **Poor social support** | 11 (20.8) | 2.3 (.59 – 9.2) | .23 | 2.4 (.57 – 10.35) | .23 | 1.9 (.40 – 8.88) | .43 |
| **Living alone** | 8 (16.3) | 1.8 (.37 – 8.30) | .48 | 0.8 (.15 – 4.92) | .85 | 2.0 (.21 – 18.16) | .55 |
| **Not in education, employment, or training (NEET)** | 13 (26.0) | 1.2 (.35 – 4.37) | .75 | 0.3 (.08 – 1.28) | .11 | .44 (.10 – 1.89) | .27 |
| **Not completed degree and not currently in university** | 24 (46.2) | 2.2 (.73 – 6.78) | .16 | 2.0 (.52 – 7.72) | .32 | 4.4 (.83 – 23.23) | .08 |
| **Personal income <£18,000 per annum** | 25 (73.5) | 0.2 (.03 – 1.11) | .07 | 1.5 (.22 – 10.04) | .68 | 1.5 (.22 – 10.04) | .68 |

### Supplement 5

Table: Sensitivity analysis – regression analysis on current informal support accessed by young adults in the whole sample and in those with disorder – both when excluding family and friends support (with IPW)

|  | Any informal support in the whole sample<br>(excluding friends and family) |  | Any informal support in those with disorder<br>(excluding friends and family) |  |
| --- | --- | --- | --- | --- |
|  | OR<br>(95% CI) | P-value | OR<br>(95% CI) | P-value |
| Any current psychiatric disorder | 4.3 (2.48-7.35) | <.001 | - | - |
| Any current depressive disorder | 2.7 (1.58-4.74) | <.001 | - | - |
| Any current anxiety disorder | 5.1 (2.92-8.83) | <.001 | - | - |
| Current comorbidity | 2.8 (1.50-5.04) | .001 | 1.1 (0.53-2.21) | .82 |
| Current self-harm/suicidal thoughts | 0.6 (0.26-1.49) | .29 | 0.29 (0.11-0.74) | .01 |
| SDQ total difficulties score | 1.1 (1.08-1.18) | <.001 | 1.1 (1.00-1.15) | .04 |
| SDQ impairment score | 1.3 (1.13-1.42) | <.001 | 1.1 (0.98-1.29) | .11 |
| Gender (female) | 0.8 (0.48-1.34) | .40 | 0.8 (0.40-1.69) | .82 |
| Age (years) | 0.9 (0.81-1.02) | .10 | 0.8 (0.67-0.94) | .01 |
| Poor social support | 1.9 (0.97-3.68) | .06 | 2.5 (1.02-6.15) | .05 |
| Living alone | 0.8 (0.36-1.80) | .60 | 0.3 (0.09-0.89) | .03 |
| Not in education, employment, or training (NEET) | 2.0 (1.03-3.75) | .04 | 0.8 (0.34-1.66) | .48 |
| Not completed degree and not currently in university | 0.7 (0.40-1.13) | .13 | 1.3 (0.62-2.59) | .51 |
| Personal income <£18,000 per annum | 2.9 (1.33-6.45) | .01 | 2.4 (0.85-6.81) | .10 |

### Supplement 6: Table: Qualitative responses on satisfaction with help received from services

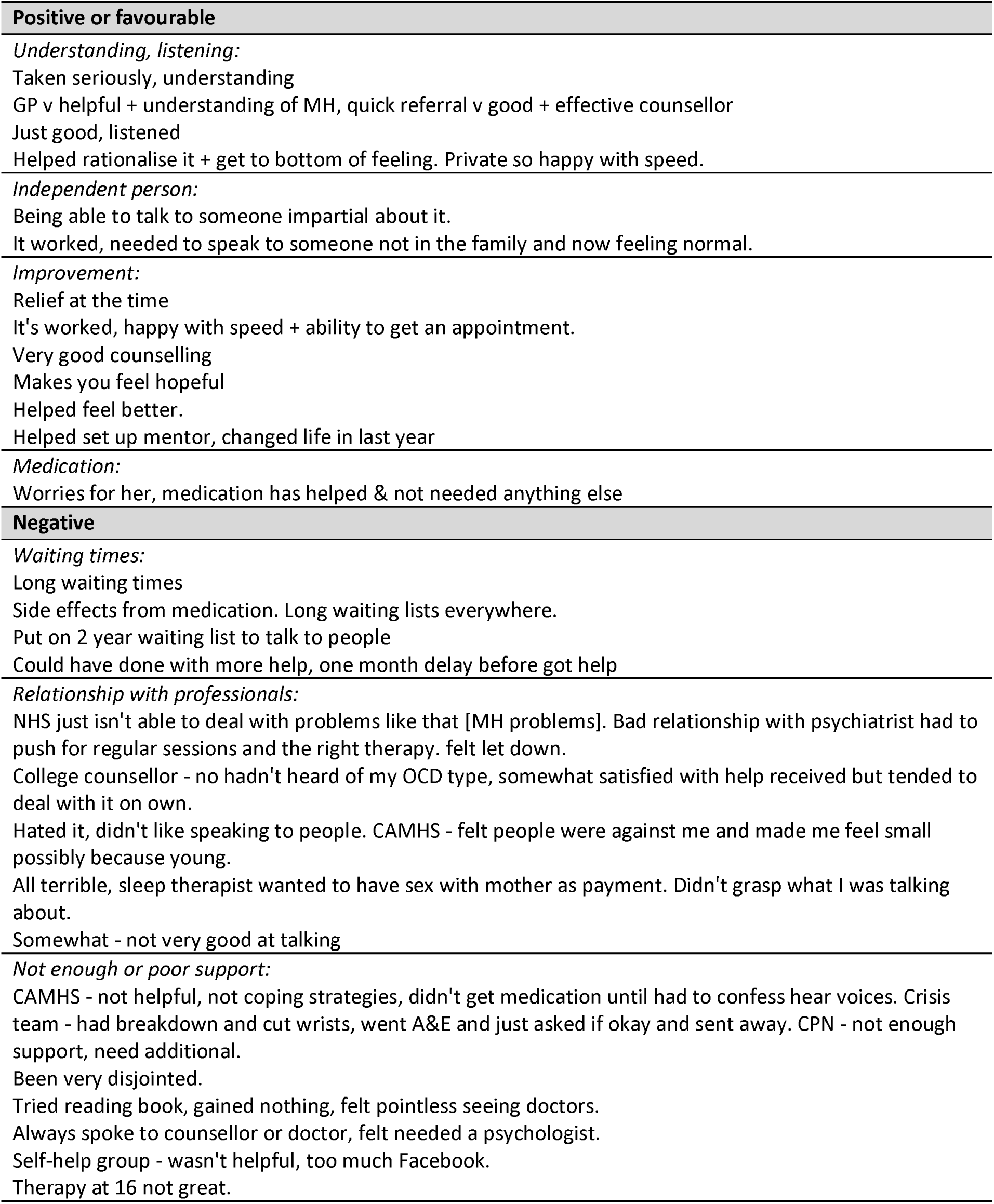
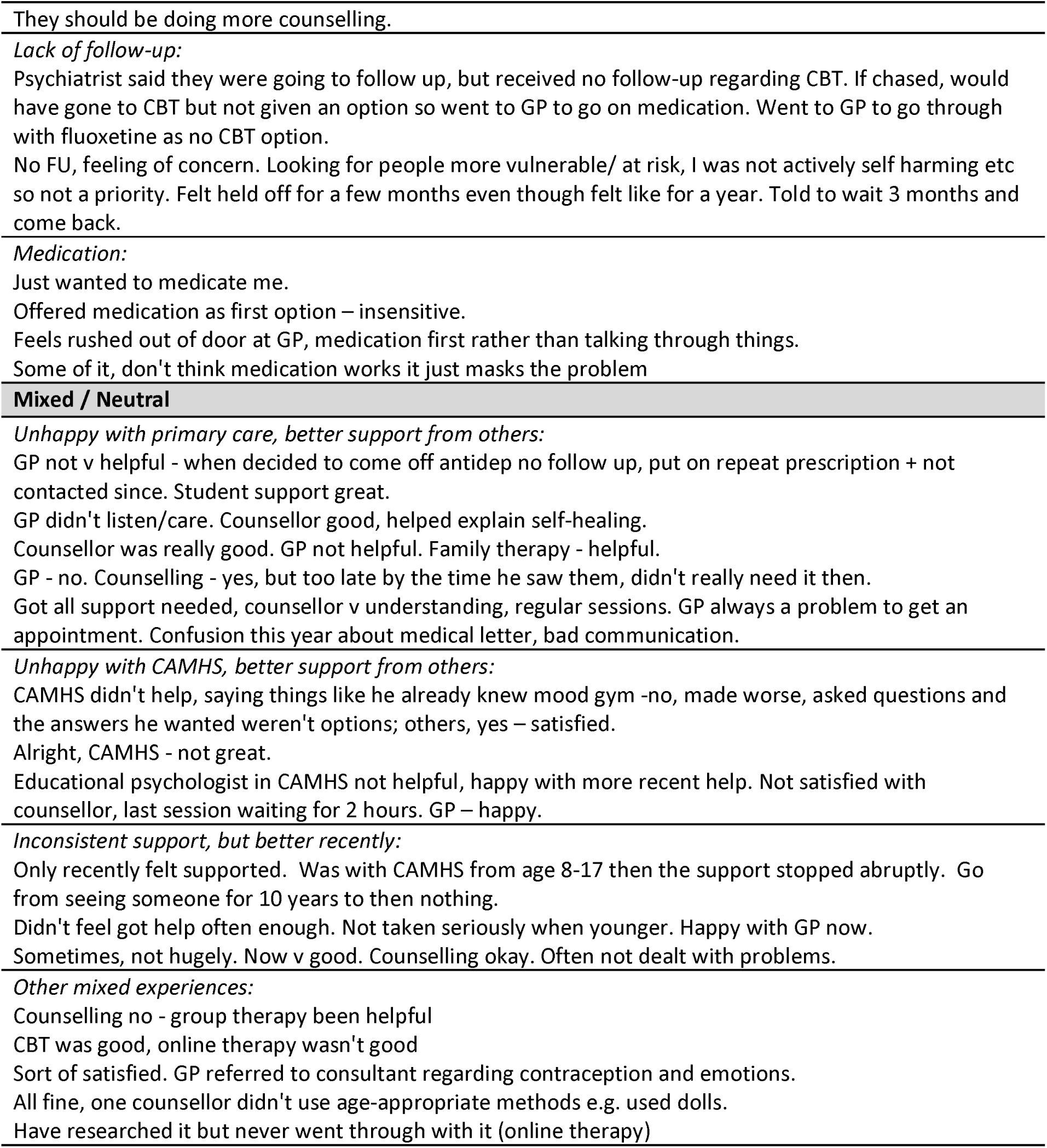
Question: If you have ever used services for help with mental health, were you satisfied with the help you received? (Yes/No) Why?

### Supplement 7 – Table: themes, subthemes, and quotes from focus group – on access to support for mental health difficulties

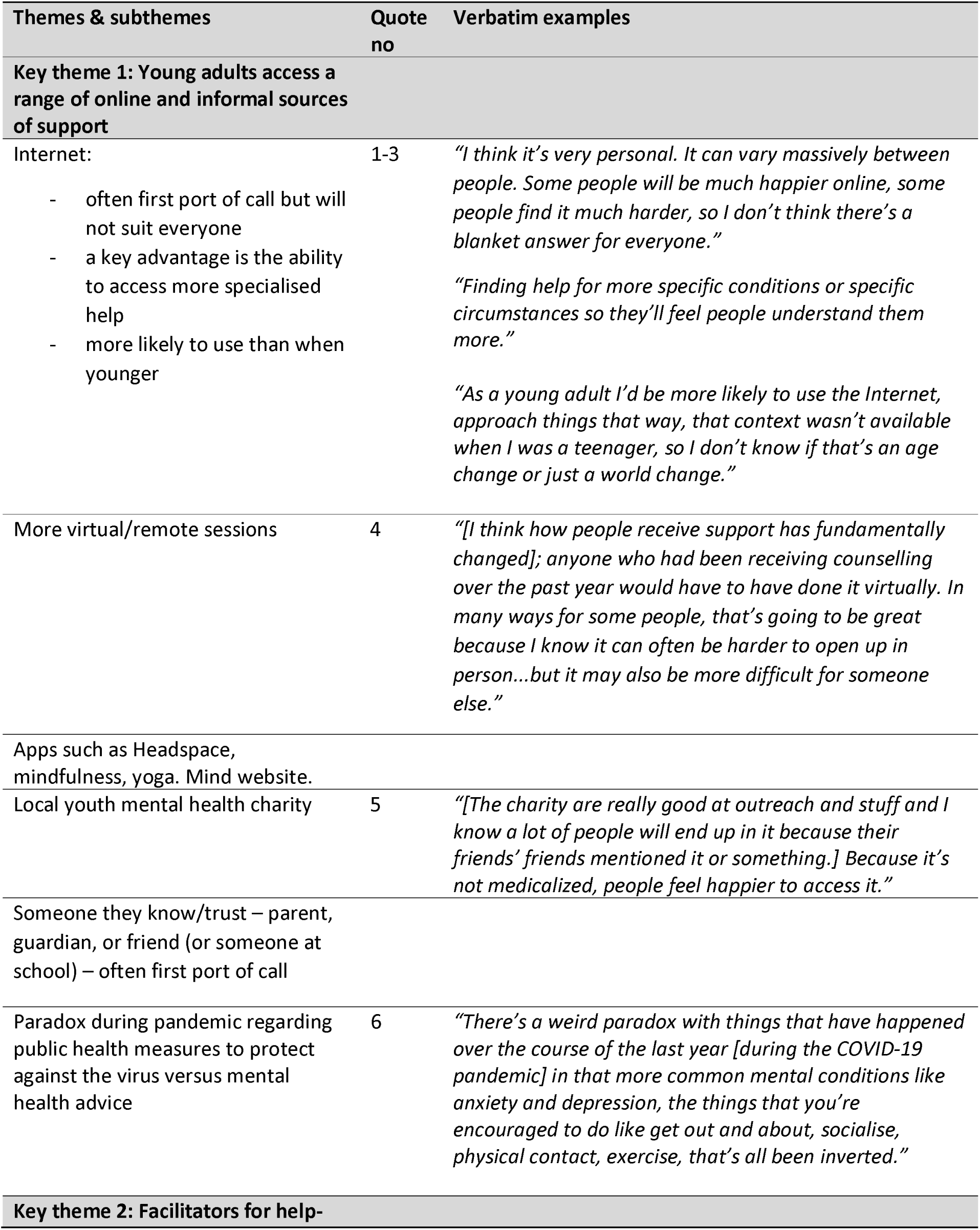
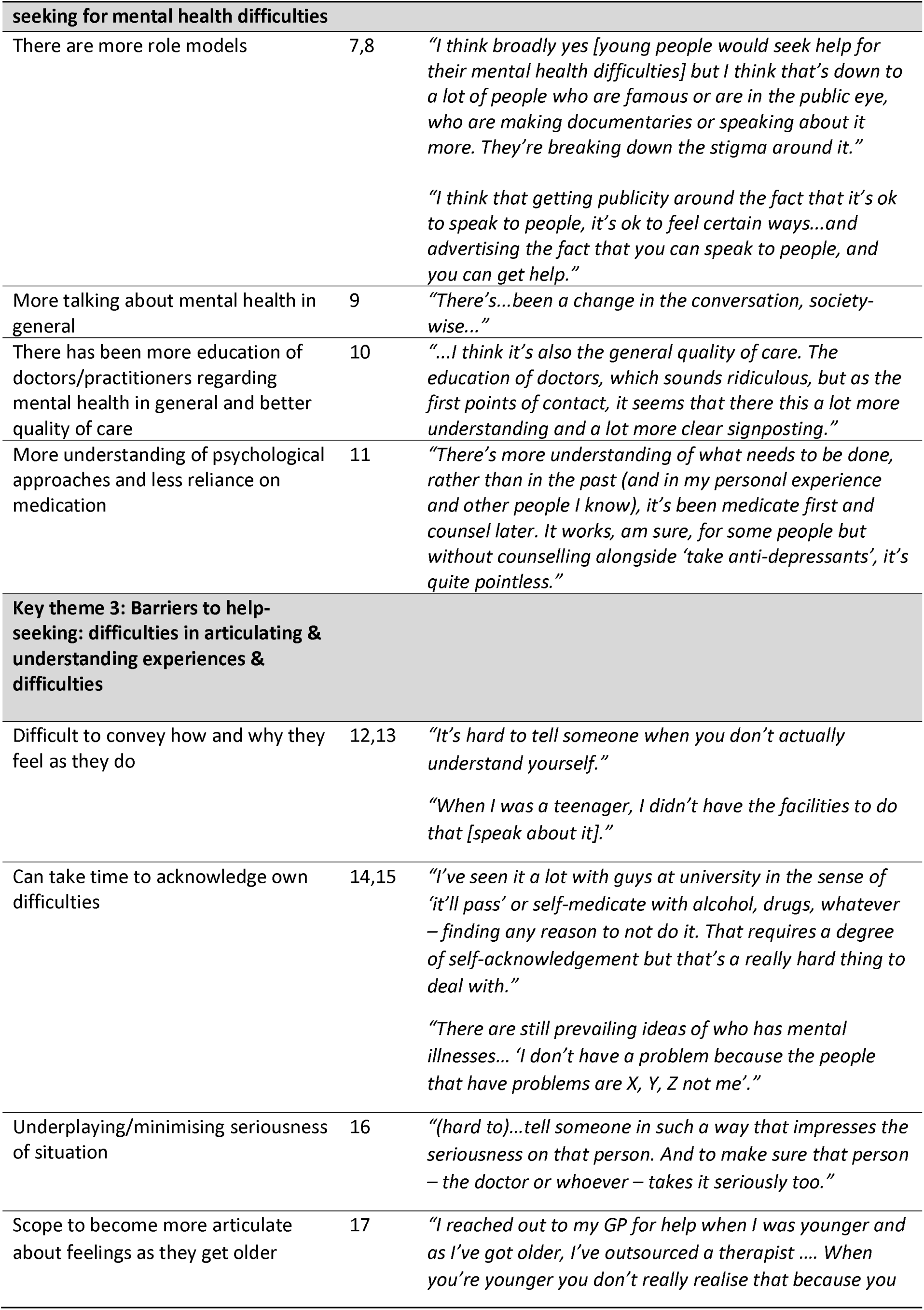
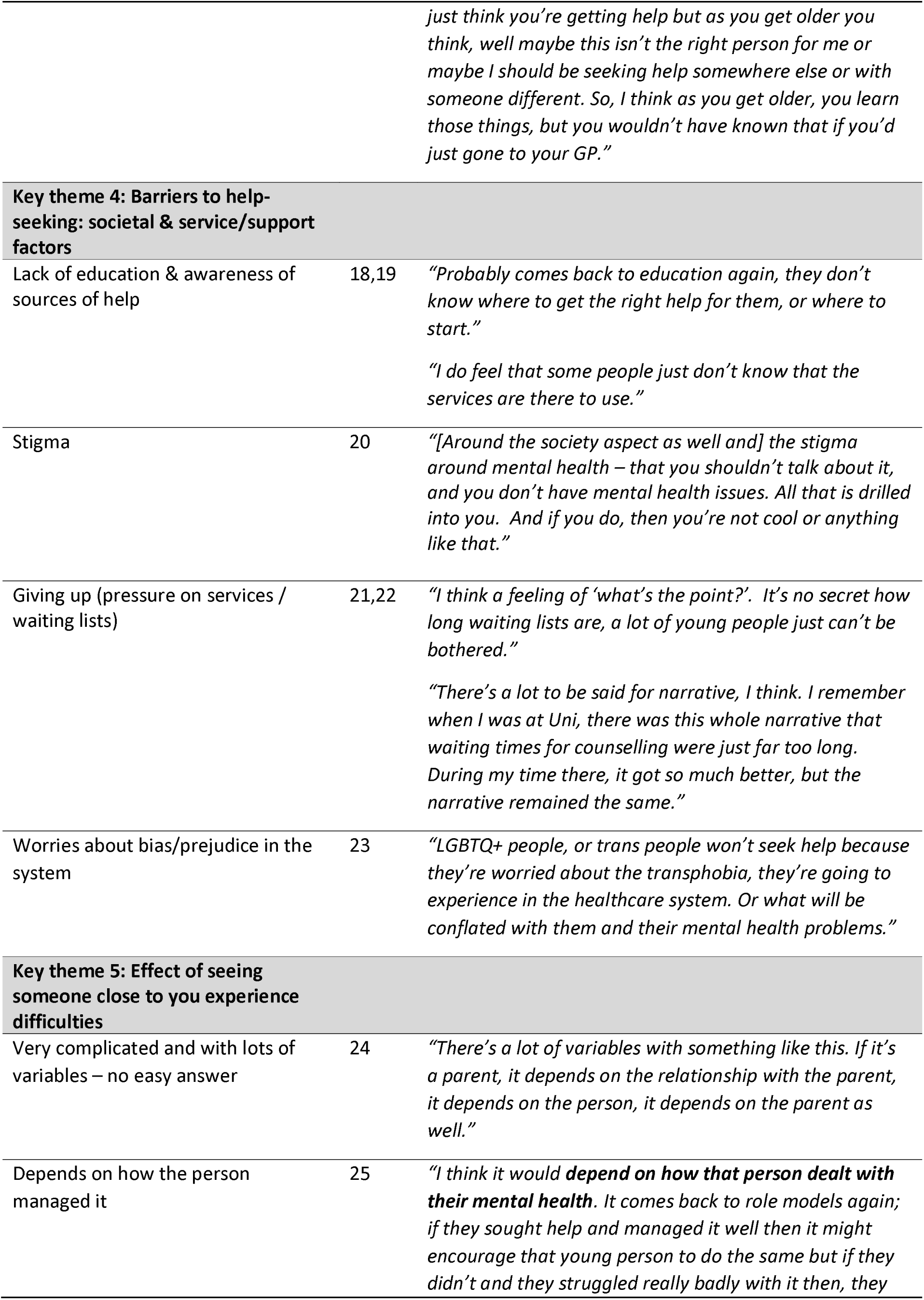
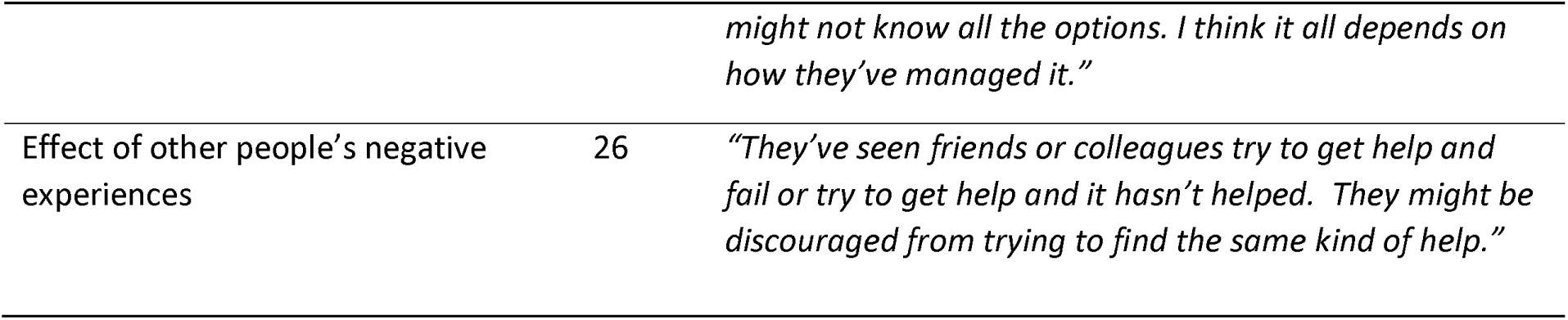

| Themes & subthemes | Quote no | Verbatim examples |
| --- | --- | --- |
| **Key theme 1: Young adults access a range of online and informal sources of support** |  |  |
| Internet:<br>- often first port of call but will not suit everyone<br>- a key advantage is the ability to access more specialised help<br>- more likely to use than when younger | 1-3 | *"I think it's very personal. It can vary massively between people. Some people will be much happier online, some people find it much harder, so I don't think there's a blanket answer for everyone."*<br>*"Finding help for more specific conditions or specific circumstances so they'll feel people understand them more."*<br>*"As a young adult I'd be more likely to use the Internet, approach things that way, that context wasn't available when I was a teenager, so I don't know if that's an age change or just a world change."* |
| More virtual/remote sessions | 4 | *"[I think how people receive support has fundamentally changed]; anyone who had been receiving counselling over the past year would have to have done it virtually. In many ways for some people, that's going to be great because I know it can often be harder to open up in person...but it may also be more difficult for someone else."* |
| Apps such as Headspace, mindfulness, yoga. Mind website. |  |  |
| Local youth mental health charity | 5 | *"[The charity are really good at outreach and stuff and I know a lot of people will end up in it because their friends' friends mentioned it or something.] Because it's not medicalized, people feel happier to access it."* |
| Someone they know/trust – parent, guardian, or friend (or someone at school) – often first port of call |  |  |
| Paradox during pandemic regarding public health measures to protect against the virus versus mental health advice | 6 | *"There's a weird paradox with things that have happened over the course of the last year [during the COVID-19 pandemic] in that more common mental conditions like anxiety and depression, the things that you're encouraged to do like get out and about, socialise, physical contact, exercise, that's all been inverted."* |
| **Key theme 2: Facilitators for help-** |  |  |

| seeking for mental health difficulties |  |  |
| --- | --- | --- |
| There are more role models | 7,8 | *"I think broadly yes [young people would seek help for their mental health difficulties] but I think that's down to a lot of people who are famous or are in the public eye, who are making documentaries or speaking about it more. They're breaking down the stigma around it."*<br>*"I think that getting publicity around the fact that it's ok to speak to people, it's ok to feel certain ways...and advertising the fact that you can speak to people, and you can get help."* |
| More talking about mental health in general | 9 | *"There's...been a change in the conversation, society-wise..."* |
| There has been more education of doctors/practitioners regarding mental health in general and better quality of care | 10 | *"...I think it's also the general quality of care. The education of doctors, which sounds ridiculous, but as the first points of contact, it seems that there this a lot more understanding and a lot more clear signposting."* |
| More understanding of psychological approaches and less reliance on medication | 11 | *"There's more understanding of what needs to be done, rather than in the past (and in my personal experience and other people I know), it's been medicate first and counsel later. It works, am sure, for some people but without counselling alongside 'take anti-depressants', it's quite pointless."* |
| **Key theme 3: Barriers to help-seeking: difficulties in articulating & understanding experiences & difficulties** |  |  |
| Difficult to convey how and why they feel as they do | 12,13 | *"It's hard to tell someone when you don't actually understand yourself."*<br>*"When I was a teenager, I didn't have the facilities to do that [speak about it]."* |
| Can take time to acknowledge own difficulties | 14,15 | *"I've seen it a lot with guys at university in the sense of 'it'll pass' or self-medicate with alcohol, drugs, whatever – finding any reason to not do it. That requires a degree of self-acknowledgement but that's a really hard thing to deal with."*<br>*"There are still prevailing ideas of who has mental illnesses... 'I don't have a problem because the people that have problems are X, Y, Z not me'."* |
| Underplaying/minimising seriousness of situation | 16 | *"(hard to)...tell someone in such a way that impresses the seriousness on that person. And to make sure that person – the doctor or whoever – takes it seriously too."* |
| Scope to become more articulate about feelings as they get older | 17 | *"I reached out to my GP for help when I was younger and as I've got older, I've outsourced a therapist .... When you're younger you don't really realise that because you* |
|  |  | *just think you're getting help but as you get older you think, well maybe this isn't the right person for me or maybe I should be seeking help somewhere else or with someone different. So, I think as you get older, you learn those things, but you wouldn't have known that if you'd just gone to your GP."* |
| **Key theme 4: Barriers to help-seeking: societal & service/support factors** |  |  |
| Lack of education & awareness of sources of help | 18,19 | *"Probably comes back to education again, they don't know where to get the right help for them, or where to start."*<br>*"I do feel that some people just don't know that the services are there to use."* |
| Stigma | 20 | *"[Around the society aspect as well and] the stigma around mental health – that you shouldn't talk about it, and you don't have mental health issues. All that is drilled into you. And if you do, then you're not cool or anything like that."* |
| Giving up (pressure on services / waiting lists) | 21,22 | *"I think a feeling of 'what's the point?'. It's no secret how long waiting lists are, a lot of young people just can't be bothered."*<br>*"There's a lot to be said for narrative, I think. I remember when I was at Uni, there was this whole narrative that waiting times for counselling were just far too long. During my time there, it got so much better, but the narrative remained the same."* |
| Worries about bias/prejudice in the system | 23 | *"LGBTQ+ people, or trans people won't seek help because they're worried about the transphobia, they're going to experience in the healthcare system. Or what will be conflated with them and their mental health problems."* |
| **Key theme 5: Effect of seeing someone close to you experience difficulties** |  |  |
| Very complicated and with lots of variables – no easy answer | 24 | *"There's a lot of variables with something like this. If it's a parent, it depends on the relationship with the parent, it depends on the person, it depends on the parent as well."* |
| Depends on how the person managed it | 25 | *"I think it would **depend on how that person dealt with their mental health**. It comes back to role models again; if they sought help and managed it well then it might encourage that young person to do the same but if they didn't and they struggled really badly with it then, they* |
|  |  | *might not know all the options. I think it all depends on how they've managed it."* |
| Effect of other people's negative experiences | 26 | *"They've seen friends or colleagues try to get help and fail or try to get help and it hasn't helped. They might be discouraged from trying to find the same kind of help."* |

